# A randomized, double-blind, placebo-controlled decentralized trial to assess sleep, health outcomes, and overall well-being in healthy adults reporting disturbed sleep, taking a melatonin-free supplement

**DOI:** 10.1101/2023.07.02.23292135

**Authors:** Antonija Kolobaric, Susan J Hewlings, Corey Bryant, Chris Colwell, Chris D’Adamo, Bernard Rosner, Jeff Chen, Emily K Pauli

## Abstract

Inadequate sleep is a global health concern. Sleep is multidimensional and complex; new multi-ingredient agents are needed. This study assessed the comparative effects of two multi-ingredient supplements on sleep relative to placebo. Adults (N=620) seeking better sleep were randomly assigned to receive 1 of 3 study products (Sleep A, Sleep B or placebo) for 4 weeks. Both active products contained federally legal hemp-derived cannabinoids, botanical oils, GABA and L-theanine. Sleep disturbance was assessed at baseline and weekly using NIH’s Patient-Reported Outcomes Measurement Information System (PROMIS™) Sleep Disturbance SF 8A survey. Anxiety, stress, pain, and well-being were assessed using validated measures at baseline and weekly. A linear mixed-effects regression model was used to assess the change in health outcome score between active product groups and the placebo. There was a significant difference in sleep disturbance, anxiety, stress, and well-being between Sleep A and placebo. There was no significant difference in any health parameter between Sleep B and placebo. Side effects were mild or moderate. There were no significant differences in the frequency of side effects between the study groups. A botanical blend containing a low concentration of THC improved sleep disturbance, anxiety, stress, and well-being in healthy individuals that reported better sleep as a primary health concern.

## 1. Introduction

Inadequate sleep has become a global public health concern, leading to greater awareness of the negative impact from a lack of sleep. Inadequate sleep increases obesity and inflammation, impairs immune and antioxidant defenses, and negatively impacts mood [1,2]. Inadequate sleep is associated with heightened emotional reactivity, reduced attention, and impaired memory and cognitive function [3]. Individuals who are sleep deprived are less productive and report a lower overall quality of life [4]. It has been suggested that shorter sleep duration alters the gut microbiome. In turn, these changes in gut microbiome may drive increases in systemic inflammation associated with metabolic syndrome and many other lifestyle related conditions [5].

It is recommended that anyone suffering with sleep disturbances consult a health professional and consider a multidimensional assessment, recognizing that an intervention that targets sleep onset may not accurately address sleep latency. Furthermore, because sleep involves multiple mechanisms, it may be advantageous to consider a multi-ingredient therapeutic approach targeting a variety of pathways to support sleep. There are many treatments aimed at improving sleep, including behavioral management, lifestyle management, exercise, diet, pharmacological interventions, and dietary supplements [6]. Many individuals experience negative side effects from prescribed medications, and choose to seek alternate solutions, such as dietary supplements and plant-based alternatives [7].

Cannabinoids are a potential plant-based alternative to prescription products for improving sleep. The cannabis plant is composed of 120 different phytocannabinoids. Delta-9 tetrahydrocannabinol (THC) and cannabidiol (CBD) are perhaps the most widely known and researched, but others like CBN are growing in popularity [8,9]. Cannabinoids produce varying effects in the human body by interacting with the endocannabinoid system (ECS), which is located throughout the brain and the central and peripheral nervous system [8]. The ECS has been suggested to modulate the circadian sleep/wake cycle. Therefore, the role of cannabinoids on sleep modulation is supported by the role of the endocannabinoid system on circadian regulation [9]. A recent systematic review, including 14 preclinical and 12 clinical studies, concluded “there are promising signs in a number of therapeutic applications that warrant additional study and there is a clear need for intensification of high-quality research into the safety and efficacy of cannabinoid therapies for treating sleep disorders….”[10]. CBD is non-intoxicating and has been shown to be safe and well-tolerated in humans, even at very high doses (e.g., 1500 mg twice daily for six days or as an acute dose of 6000 mg [11]. Cannabinol (CBN) is a by-product of THC and is found in small amounts in the cannabis plant. CBN has gained consumer interest as an ingredient to benefit sleep [12,13]. However, research related to CBN and sleep is limited, and the majority of it dates back to the 1970s and 80s [14].

Other non-pharmacological alternatives have been investigated. For example, γ-aminobutyric acid (GABA) is a non-protein amino acid, well known for its role as an inhibitory neurotransmitter in the central nervous system, promoting relaxation and sleep [15,16]. Additionally, γ-Glutamylethylamine, also known as L-theanine, is a non-protein amino acid found in green tea, often used to improve sleep and modify stress. It is thought to increase expression of dopamine and serotonin in the brain by increasing GABA levels [17,18]. It has been suggested that these compounds provide synergistic benefits. In mice, GABA/L-theanine mixture (100/20 mg/kg) demonstrated a decrease in sleep latency (20.7 and 14.9%) and an increase in sleep duration (87.3 and 26.8%) compared to GABA or L-theanine alone [18].

Botanical essential oils are also gaining popularity for their effectiveness in improving sleep. Hops is an herbaceous perennial plant belonging to the class Magnoliopsida, subclass Hamamelididae, order Urticales, family Cannabaceae, genus Humulus. It has been known for centuries for its many health benefits, including its sedative effects [19]. Several studies support the sedative properties of hops oil [20,21]. The sedative effect is thought to occur due to a degradation product, 2-methyl-3-buten-2-ol, that increases GABA [22]. Alone, the amount of any one botanical is most likely too low to lead to any significant benefit, but it is believed that the synergy with other phytocompounds produces the effect [23,24].

Another popular oil often used together with hops, is Valeriana L., a group of perennial herbs belonging to the family Caprifoliaceae. It has been used for centuries around the globe to improve sleep. The roots and rhizomes are most often used for medicinal benefits. There are more than 200 species of Valeriana, with Valeriana officinalis L used most often in US products. In mice, the essential oil was determined to be the active part of the plant. The compounds within the oils could act synergistically on GABA receptors to increase GABA release and inhibit uptake [25-27].

Though each of the botanicals discussed thus far has shown promise for improving sleep, it is intriguing to consider the concept of synergy, especially because sleep is multidimensional and complex, thus warranting a multi-ingredient approach. The discussion of botanical synergy is not new. In recent years, it has been commonly called “the entourage effect” when discussed in reference to the potential therapeutic effects of the synergy between phytocannabinoids and the many other compounds found in the cannabis plant [28]. Demonstrated by several review papers, the application of botanical synergy extends beyond the cannabis plant to other botanicals [29,30]. It has been suggested that synergy among the cannabinoid compounds may enhance the effectiveness of THC, thus allowing a lower effective dose, minimizing the psychoactive effects of THC while maintaining the benefits [31]. Whether compounds from other botanicals would provide a similar synergistic effect is unknown.

The aim of this study was to assess the effects of two different softgel dietary supplements, one with lower THC and higher levels of other botanicals (Sleep A), and one with higher THC and lower levels of other botanicals (Sleep B), on sleep disturbance relative to placebo.

## 2. Materials and Methods

This study, Radicle™ Rest, was a randomized, double-blind, placebo-controlled, parallel group, virtual trial, designed to assess the effects of health and wellness products on sleep, anxiety, stress, pain, and overall health-related quality of life. Participants were not required to attend in-person visits. All data were collected via online surveys which participants accessed via participant specific hyperlinks sent to them at scheduled times through their preferred means of communication (email or SMS text). Participants were recruited online from across the US through social media, Radicle Science’s electronic mailing list, and a third-party consumer network with nationwide representation. Recruitment emails containing links to the study screener were sent to those within the Radicle Science mailing list and consumer network, while social media advertisements led to a study landing page with a link to the study screener. Participants were eligible if they were 21 years old or older, resided in the United States, expressed a desire for better sleep, and ranked their desire for better sleep as a primary reason for taking a dietary supplement. Individuals were excluded if they were pregnant or breastfeeding, or taking medications that posed a health risk when used in conjunction with any of the study product ingredients. Eligible individuals were directed to a secure online portal to provide informed consent. Participants indicated their consent electronically by signing the informed consent form and were sent a digital copy of the electronic consent. Eligible individuals were advised to consult with their healthcare provider before participating if they had a diagnosed medical condition, were on any prescription medication or supplements, or had any upcoming medical procedures planned. Immediately following informed consent, participants completed an intake survey which collected basic demographic information, health behaviors, and experienced sleep quality.

Recruits who consented to participate and completed intake were randomized to one of three study arms (see below for details on randomization): Sleep A, Sleep B, or Placebo (Figure 1).

**Figure 1.**
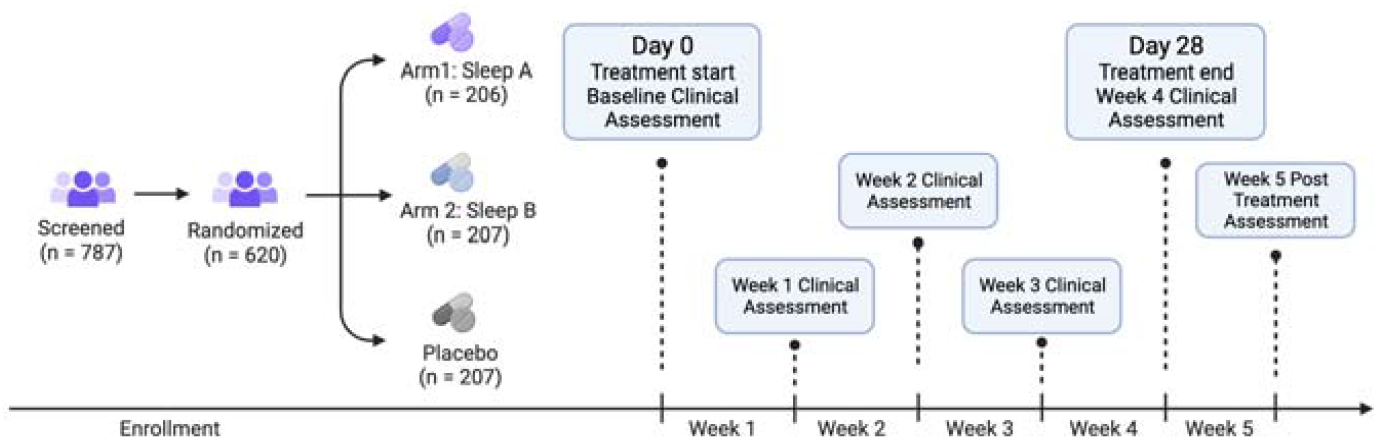
Study Flow Diagram. Eligible participants were enrolled in the study and randomized into one of three groups: Sleep A, Sleep B, or placebo. We collected baseline clinical measures before participants started using their study product. Participants used study product for 4 weeks total. Clinical and other measures were collected at the end each week as well as 1 week post study product use.

Participants were sent a 4-week supply of their study product in the mail, along with the product insert, detailing instructions for study participation. All study products were provided by the partnering manufacturer and analyzed at an independent laboratory to ensure active ingredient identification, safety, and potency. Participants were instructed to take one softgel 30 minutes before bedtime, and informed that they could escalate to a maximum of 4 softgels per day as needed throughout the study duration. Participants were directed to wait 5 days before increasing the number of softgels taken. The study product formulations are proprietary to the manufacturer but both Sleep A and B formulations contained the same amount of CBD, CBN, and L-theanine; Sleep A formula contained lower amounts of THC and higher amounts of GABA, hops oil and valerian oil relative to the Sleep B formula. The study was double-blind; neither the participants nor those who collected and analyzed the data were aware of the product participan s received until the conclusion of the study. The study was conducted from Oct 2022 to Dec 2022

For 5 weeks following the study initiation and baseline (4 weeks taking the study product and 1 week after finishing the study product), participants were asked to complete online surveys, which they accessed via unique hyperlinks sent at scheduled times via email or text. During the baseline week, participants completed health outcome assessments of their sleep, feelings of anxiety, stress, pain, and overall well-being, using validated, patient reported outcome measures (Table 1).

**Table 1.**
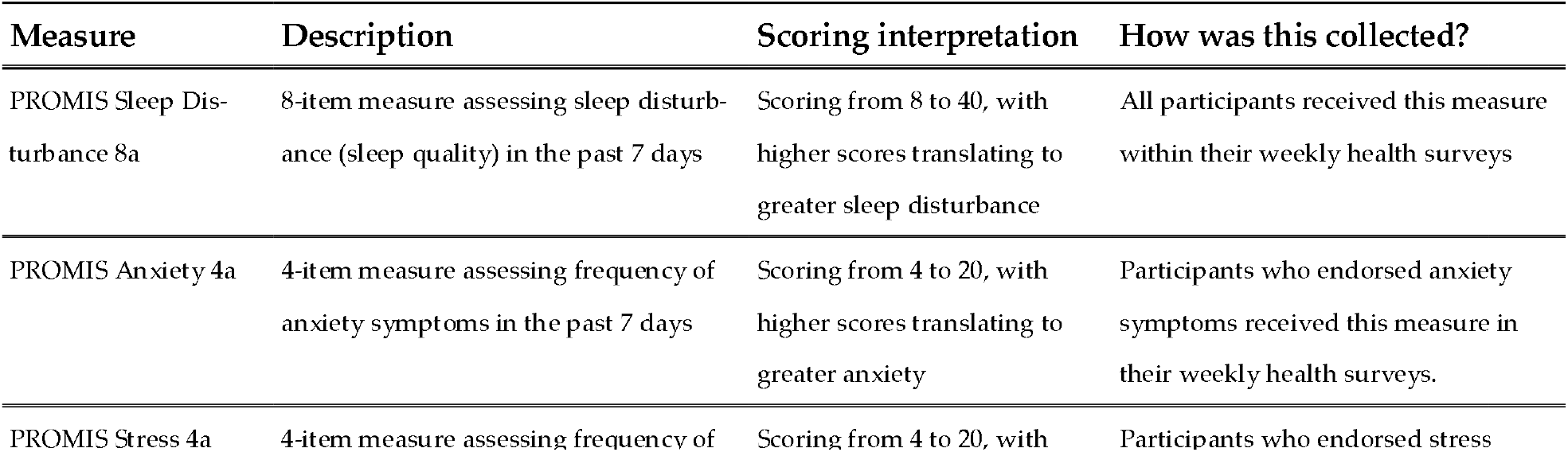
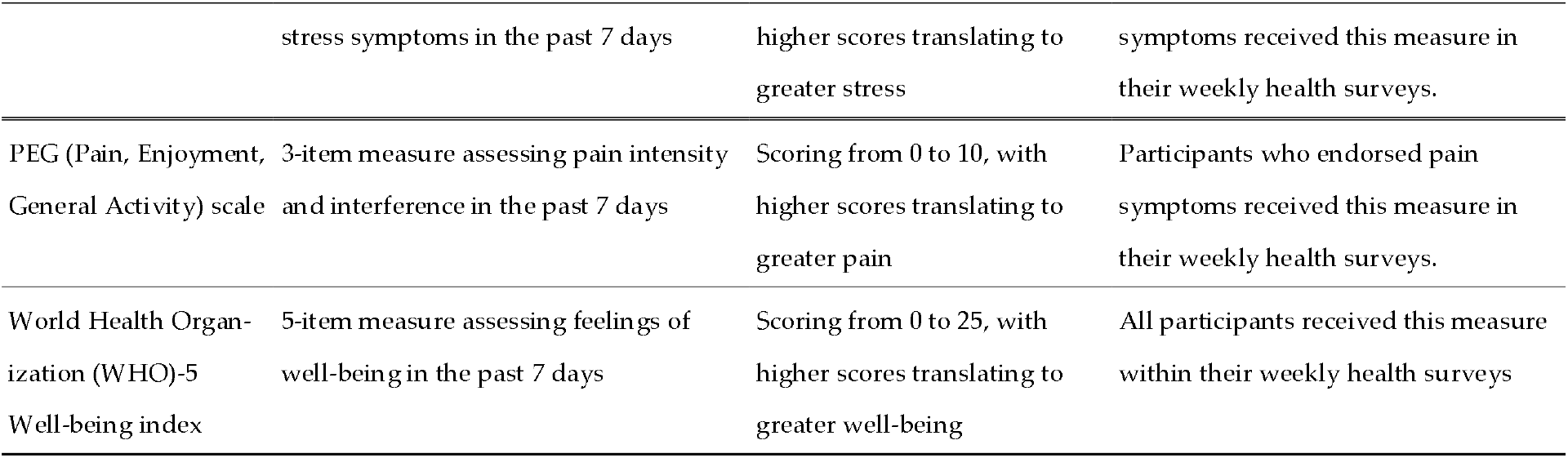
Validated measures for key outcomes used in Radicle Rest study.

| Measure | Description | Scoring interpretation | How was this collected? |
| --- | --- | --- | --- |
| PROMIS Sleep Disturbance 8a | 8-item measure assessing sleep disturbance (sleep quality) in the past 7 days | Scoring from 8 to 40, with higher scores translating to greater sleep disturbance | All participants received this measure within their weekly health surveys |
| PROMIS Anxiety 4a | 4-item measure assessing frequency of anxiety symptoms in the past 7 days | Scoring from 4 to 20, with higher scores translating to greater anxiety | Participants who endorsed anxiety symptoms received this measure in their weekly health surveys. |
| PROMIS Stress 4a | 4-item measure assessing frequency of | Scoring from 4 to 20, with | Participants who endorsed stress |

|  | stress symptoms in the past 7 days | higher scores translating to greater stress | symptoms received this measure in their weekly health surveys. |
| --- | --- | --- | --- |
| PEG (Pain, Enjoyment, General Activity) scale | 3-item measure assessing pain intensity and interference in the past 7 days | Scoring from 0 to 10, with higher scores translating to greater pain | Participants who endorsed pain symptoms received this measure in their weekly health surveys. |
| World Health Organization (WHO)-5 Well-being index | 5-item measure assessing feelings of well-being in the past 7 days | Scoring from 0 to 25, with higher scores translating to greater well-being | All participants received this measure within their weekly health surveys |

Throughout the study duration, participants received a health survey asking them to report their study product usage for the week and health outcome assessments for their sleep disturbance, feelings of anxiety, stress, pain and overall well-being from the past week using the same validated health measures used at baseline. In every study survey, following receipt of their product, participants were also prompted to report any side effects, and were encouraged to contact the research team directly if they experienced side effects at any point.

Sterling Independent Review Board (SIRB) approved the study [10147]. The master protocol Radicle Rest™ was registered on ClinicalTrials.gov[Identifier: NCT05511818]. It should be noted that the protocol registered is not this specific study but rather is the templated study protocol utilized for all Radicle Rest studies.

### 2.1. Randomization

Participants were randomly assigned to one of the three study product arms, with an equal chance of being assigned to each group (1:1:1 ratio). Prior to randomization, participants were stratified by their assigned sex at birth (male, female) then randomized to one of the study arms using the randomizer with evenly presented elements in the Qualtrics® XM platform.

### 2.2. Outcomes

Radicle™ Rest is a templated trial protocol incorporating validated assessments which has received overall IRB approval that is then applied to each individual study. The study design and assessments used do not change from study to study, only the actives and placebos change. The primary outcomes were change in the PROMIS™ Sleep Disturbance 8A scale [32] as well as the odds of achieving a minimal clinically important difference (MCID). MCID was defined as a reduction which is greater than or equal to one-half the standard deviation of the baseline score [33]. The MCID standard deviation criterion was calculated by study arm. Secondary outcomes included changes in anxiety, stress, pain, and overall wellbeing. Secondary outcomes were assessed using PROMIS™ Anxiety 4a, PROMIS™ Stress 4a, PEG (Pain, Enjoyment, General Activity), and World Health Organization (WHO)-5 Well-being index.

### 2.3. Safety

The frequency of spontaneously reported side effects and their severity were assessed. Severity was determined based on reported utilization of medical services in response to the side effects according to the following grading schema based on the Common Terminology Criteria for Adverse Events (CTCAE; v5.0 USDHHS): mild: no intervention (medication or medical advice) needed; moderate: a medication was taken due to the side effect or a participant sought medical advice from their HCP, urgent clinic or ED; severe: the side effect was medically significant but not life-threatening and/or the participant was admitted to the hospital for care and attention; life threatening: immediate medical intervention required and the participant was hospitalized, placed in the intensive care unit due the side effect, and/or suffered long-lasting negative effects as a result of the side effect.

### 2.4. Covariates

Prior to analysis, we collapsed three demographic variables, including race, education, and ethnicity. Race was recoded as white (including participants who identified as white), non-white (including participants who identified as Black, Multi-racial, Asian, Unknown, Prefer not to say, Some other race, or American Indian or Alaska Native), and Native Hawaiian or Pacific Islander (including participants who identified as Native Hawaiian or Pacific Islander). Education was recoded as college degree (including participants who have a bachelor’s or associate degree, and masters or professional degree), and No college degree (including participants who selected prefer not to say, less than high school, trade/technical/vocational degree, high school diploma no college, and some college no degree). Ethnicity was recoded to Hispanic (including participants who selected Yes) and Non-Hispanic (including participants who selected no, or prefer not to say). We adjusted for baseline demographics, including age, recoded race, recoded ethnicity, recoded education level, sex assigned at birth (male, female), and body mass index (BMI; calculated through self-reported height and weight).

### 2.5. Power analysis

A power analysis was conducted to ensure sufficient power to detect a significant difference in the change in the PROMIS™ Sleep Disturbance 8A scale for each study product arm relative to placebo. A sample size of 190 for each study group would yield 90% power to find a difference in mean change between each study product arm versus the placebo arm at a two-sided p value of 0.05 corrected for multiple comparisons (Bonferroni). Recruiting up to 300 participants per study arm would allow us to maintain adequate sample size under anticipated attrition levels (45%).

### 2.6. Statistical analysis

A linear, mixed-effects regression model was used to assess the differences in the change in the variables of interest between each active product arm versus placebo. The parameter “na.action = na.omit” was set for each model, meaning that participants were excluded only from those models for which they did not have available data. All models were fit using an unstructured covariance matrix with a random-intercept at the individual level, and a random-slope and intercept at the study week level. The models tested the difference in the interaction between product arm and study week for active arm versus placebo, controlling for sex, age, race, ethnicity, and BMI. Post hoc Bonferroni-adjusted pairwise comparisons were used to assess the differences in the odds of achieving a MCID for sleep between each active product arm placebo, controlling for sex, age, race, ethnicity, education, and BMI.

### 2.7. Software

The Python programming language, version 3.95 (packages: pandas, version 1.4.3, and numpy, version 1.20.2) were used for data processing. R, version 4.2.3 (packages: nlme, version 3.1-162, marginal effects, version 0.11.1, and tidyverse, version 2.0.0) was used to conduct the statistical analyses, and package table one version 0.13.2 was used to create table one.

## 3. Results

### 3.1. Particpants

Subsubsection The study population reflects well the population of US consumers that elect to purchase health and wellness supplements. At baseline, 90% of participants reported that they suffered from stress and 76% reported that they suffered from sleep disturbances most (63.6% reported mild or moderate sleep disturbances). In this study, 66% of participants were female, 34% were male and 80% identified their race as white. After stratification, 206 participants were randomly assigned to Sleep A, 207 to Sleep B, and 207 to Placebo. The groups did not significantly differ on any demographic or outcomes variables at baseline (Table 2).

**Table 2.**
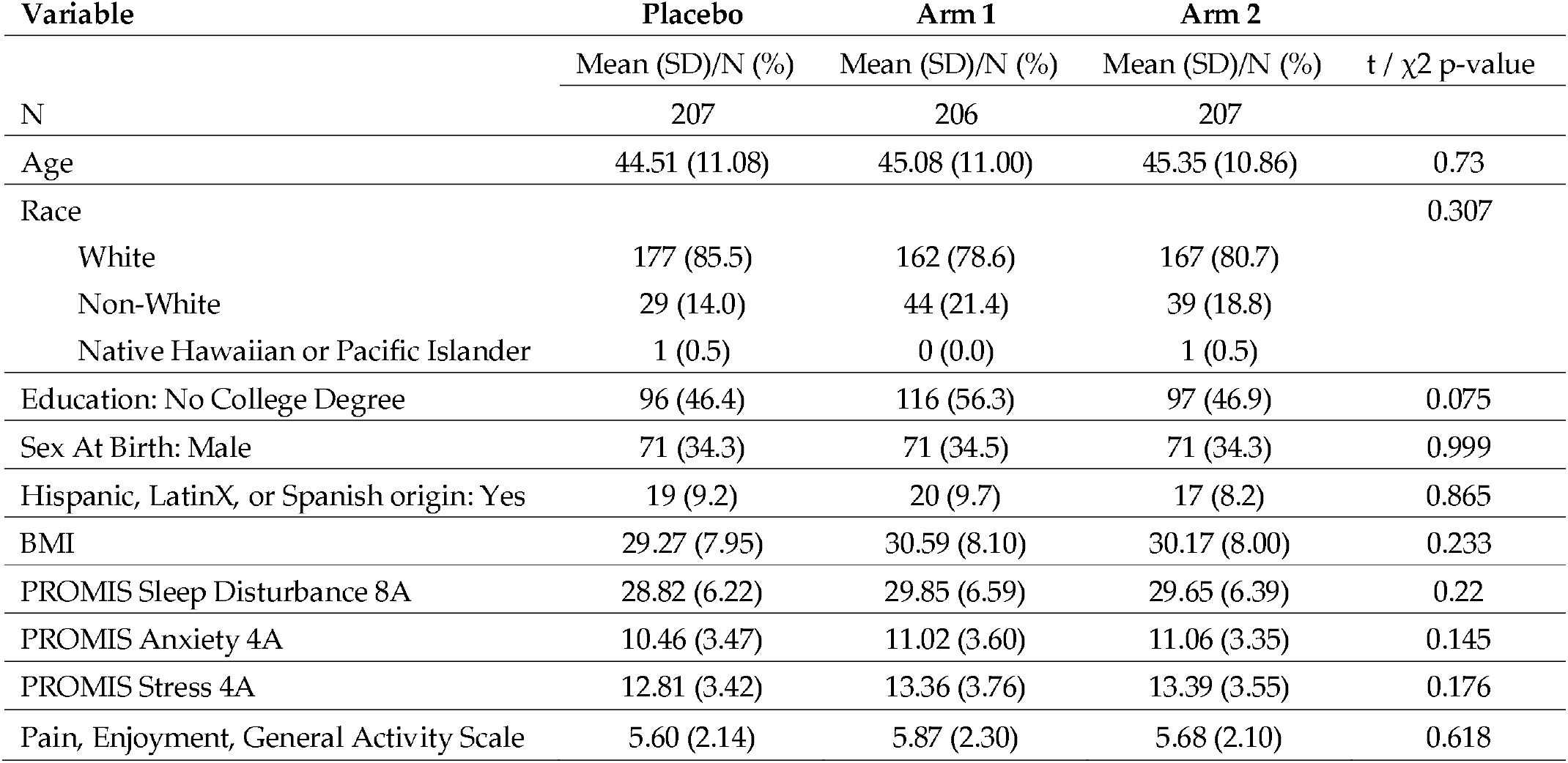
Participant sample summary at baseline.

### 3.2. Sleep Quality

The interaction between Study Week and Sleep A (Arm 1) showed a significant negative association with sleep disturbance (β = -0.639, p = 0.0027). This indicates that the effect of Study Week on sleep disturbance differed between the treatment groups, with participants in Sleep A (Arm 1) experiencing a greater reduction in sleep disturbance over time compared to the placebo group. Education demonstrated a significant positive association with sleep disturbance (β = 1.846, p < 0.0001), suggesting that individuals without a college degree reported higher levels of sleep disturbance. BMI exhibited a significant positive association with sleep disturbance (β = 0.070, p = 0.0163), indicating that higher BMI was associated with higher levels of sleep disturbance (Figure 2, Table 3). We did not observe any significant differences in the likelihood of achieving a minimum clinically important difference (MCID) in between Sleep A (estimate = 1.33, 95% CI[0.85, 1.81], p = 0.242) or or Sleep B (estimate = 1.23, 95% CI[0.79, 1.67], p = 0.477) and placebo (42.2 %). MCID is defined as a change of one half the standard deviation of the baseline score.

**Table 3.**
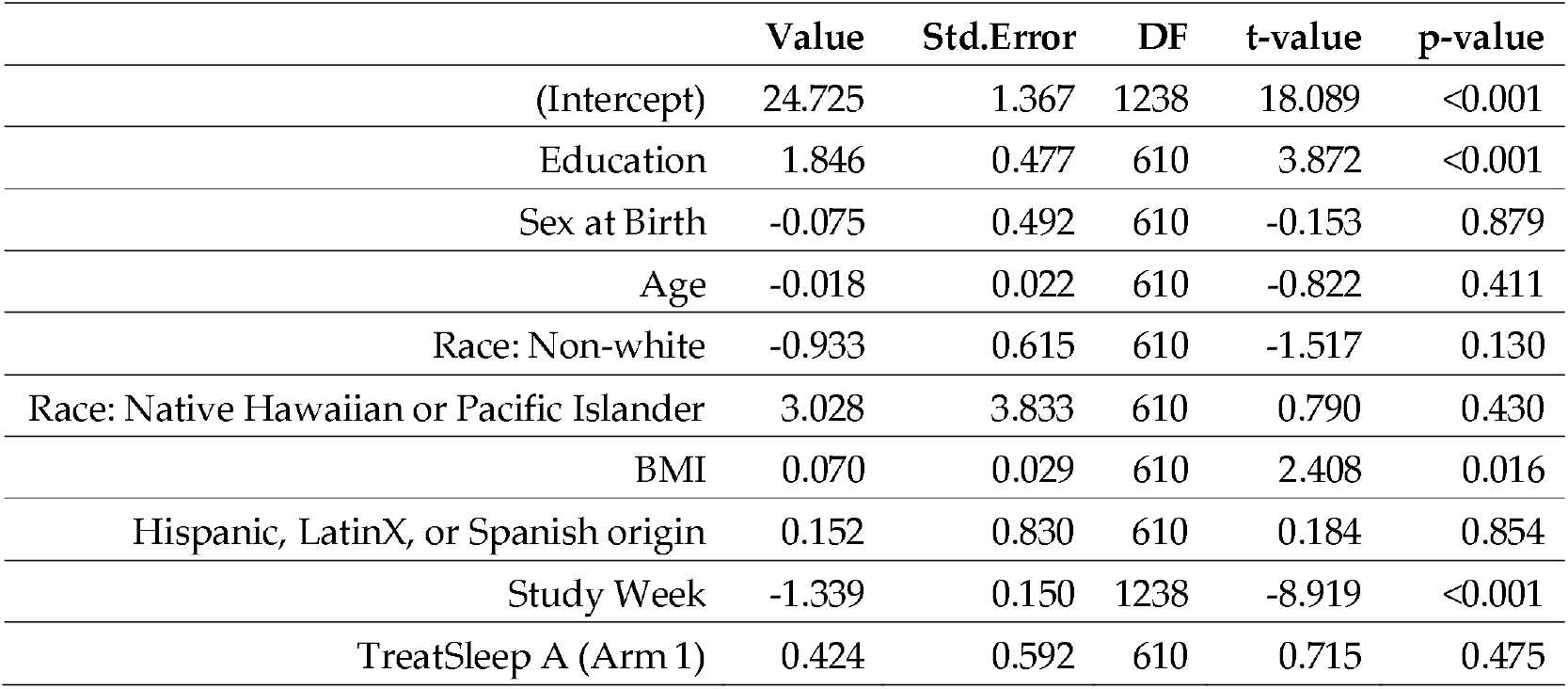
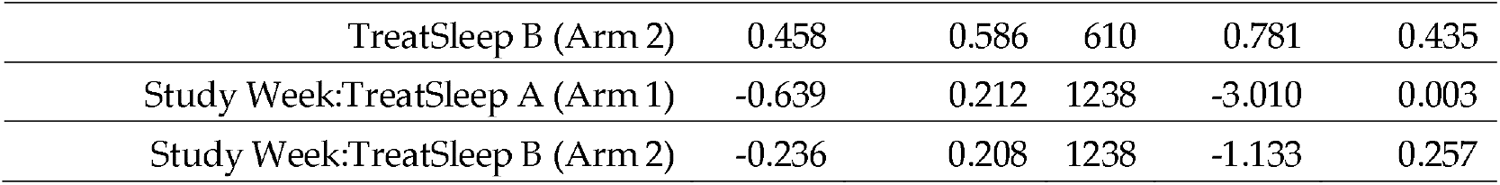
Significant Factors Associated with Sleep Disturbance: Results from a Linear Mixed Effects Regression Model. Summary of significant variables and their associations with sleep disturbance based on a linear mixed-effects regression model. The model was used to assess the differences in the change in the variables of interest between each active product arm versus placebo. The table presents the beta coefficients (β), standard errors (Std.Error), degrees of freedom (DF), t-values, and p-values for each variable. Higher values indicate a stronger positive association with sleep disturbance, while lower values indicate a stronger negative association.

**Figure 2.**
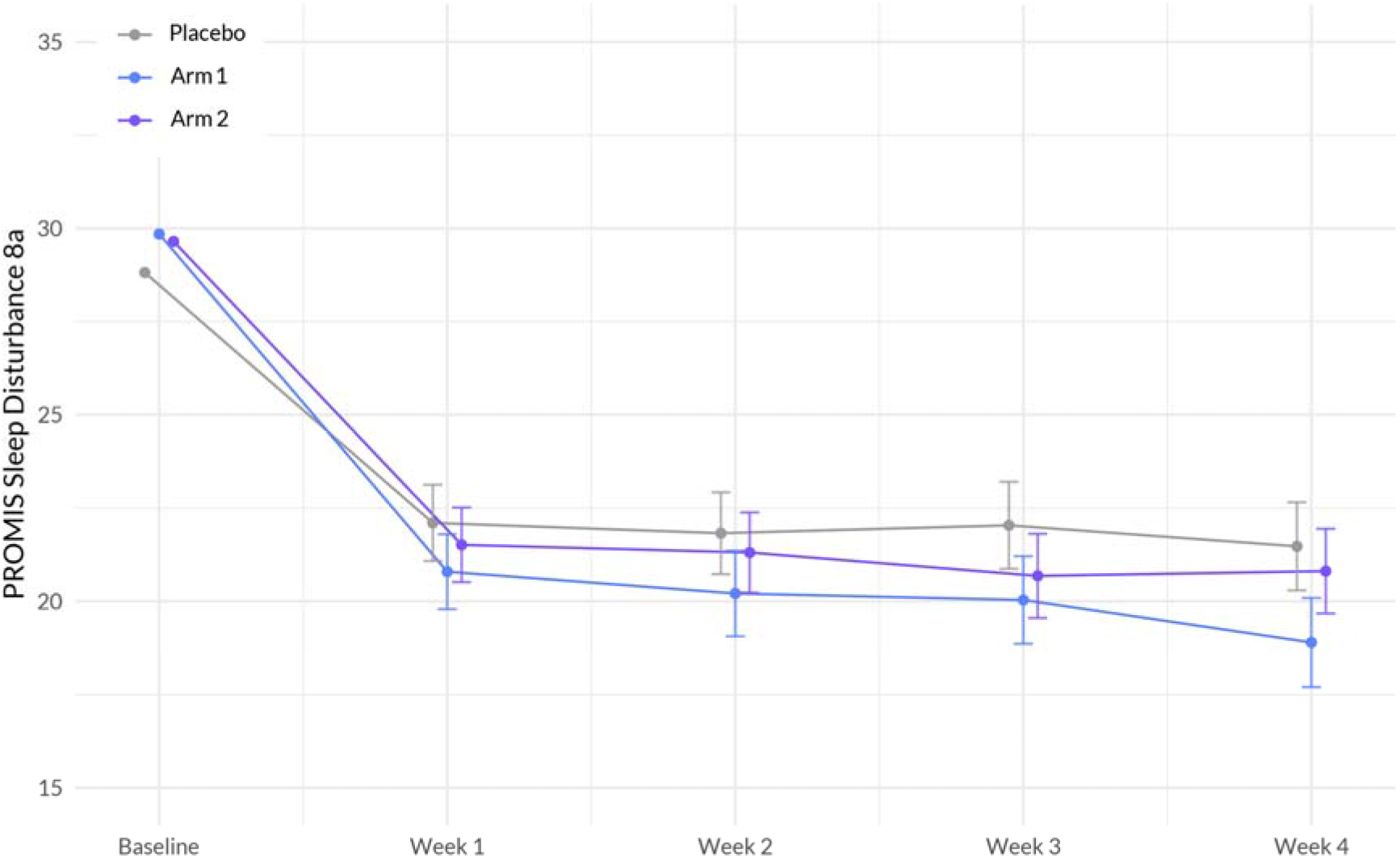
Interaction between Treatment and Week on PROMIS Sleep Disturbance 8a score. The plot illustrates the interaction between treatment (Sleep A; Arm 1 and Sleep B; Arm 2) and week on the sleep disturbance scale, based on a linear mixed-effects model. The x-axis represents the weeks of the study, while the y-axis represents the outcome scale. The lines represent the trajectories of sleep disturbance for each treatment arm over time. The plot highlights the nature of treatment effects on sleep quality, as captured by the linear mixed-effects model, allowing for the incorporation of random effects and accounting for within-subject correlations.

### 3.3. Sleep Quality

The analysis revealed several significant associations with anxiety. First, there was a significant negative interaction between Study Week and Sleep A (Arm 1) (β = -0.258, p = 0.041), indicating that as the study progressed, participants using Sleep A were more likely to experience a decrease in anxiety compared to participants using placebo (Figure 3, Table 4). Additionally, education showed a significant positive association with anxiety (β = 0.744, p = 0.005), suggesting that individuals with a higher level of education experienced higher levels of anxiety. Lastly, age demonstrated a significant negative association with anxiety (β = -0.035, p = 0.005), indicating that as age increased, anxiety levels tended to decrease.

**Table 4.** Significant Factors Associated with Anxiety: Results from a Linear Mixed Effects Regression Model. Summary of significant variables and their associations with anxiety based on a linear mixed-effects regression model. The model was used to assess the differences in the change in the variables of interest between each active product arm versus placebo. The table presents the beta coefficients (β), standard errors (Std.Error), degrees of freedom (DF), t-values, and p-values for each variable. Higher values indicate a stronger positive association with anxiety, while lower values indicate a stronger negative association.

|  | Value | Std.Error | DF | t-value | p-value |
| --- | --- | --- | --- | --- | --- |
| (Intercept) | 11.007 | 0.764 | 773 | 14.415 | <0.001 |
| Education: No College Degree [Ref: College Degree] | 0.744 | 0.266 | 610 | 2.795 | 0.005 |
| Sex at Birth: Male [Ref: Female] | -0.398 | 0.276 | 610 | -1.439 | 0.151 |
| Age | -0.035 | 0.012 | 610 | -2.855 | 0.005 |
| Race: No-white [Ref: white] | -0.027 | 0.345 | 610 | -0.078 | 0.938 |
| Race: Native Hawaiian or Pacific Islander [Ref: white] | 0.667 | 2.258 | 610 | 0.295 | 0.768 |
| BMI | 0.021 | 0.016 | 610 | 1.264 | 0.207 |
| Hispanic, LatinX, or Spanish origin: Yes [Ref: No] | 0.196 | 0.452 | 610 | 0.434 | 0.665 |
| Study Week | -0.445 | 0.087 | 773 | -5.126 | <0.001 |
| Sleep A (Arm 1) | 0.388 | 0.329 | 610 | 1.179 | 0.239 |
| Sleep B (Arm 2) | 0.577 | 0.326 | 610 | 1.769 | 0.078 |
| Study Week: Sleep A (Arm 1) | -0.259 | 0.127 | 773 | -2.044 | 0.041 |
| Study Week: Sleep B (Arm 2) | -0.066 | 0.123 | 773 | -0.537 | 0.592 |

**Figure 3.**
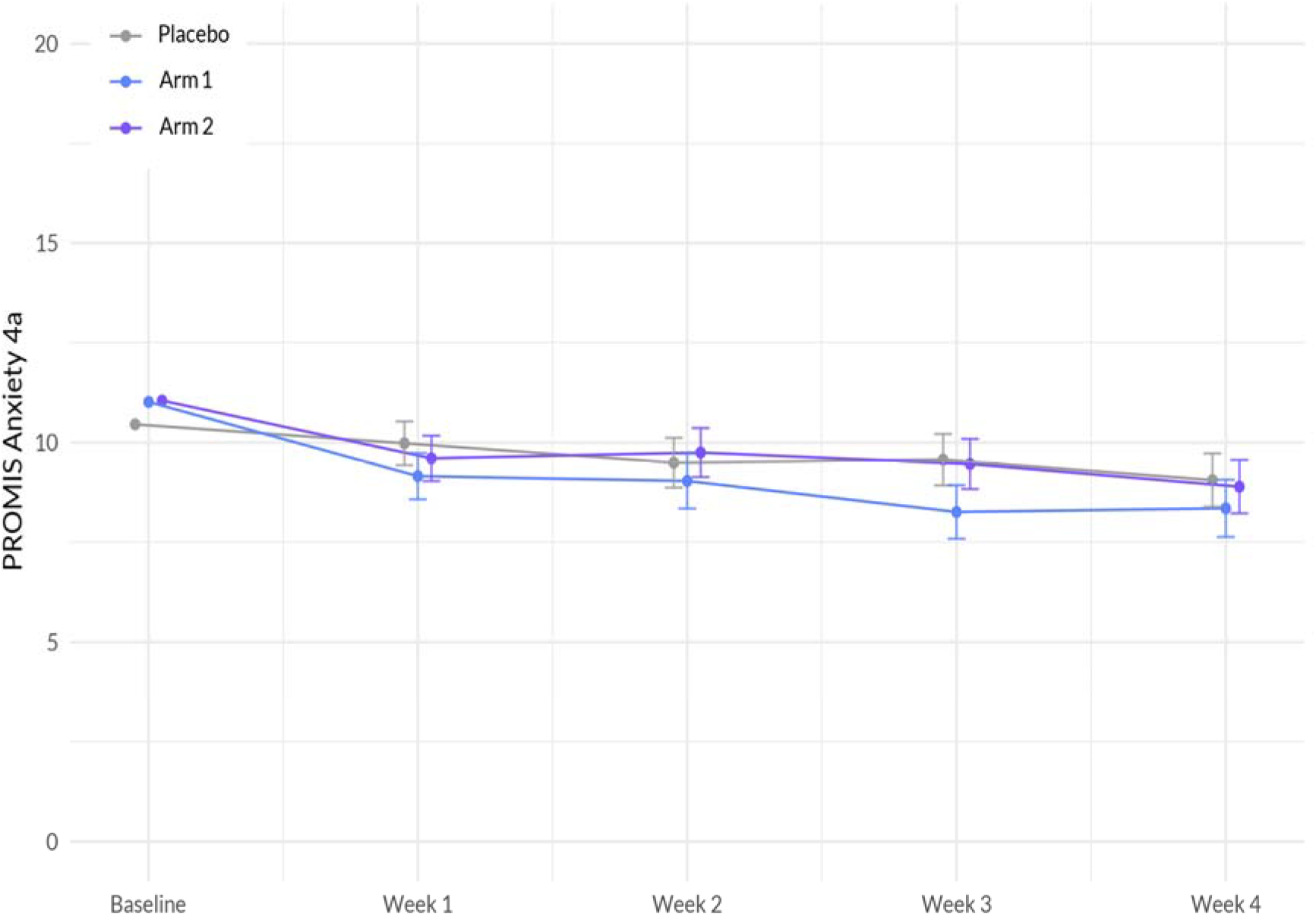
Interaction between Treatment and Week on PROMIS Anxiety 4a score. The plot illustrates the interaction between treatment (Sleep A; Arm 1 and Sleep B; Arm 2) and week on the anxiety scale, based on a linear mixed-effects model. The x-axis represents the weeks of the study, while the y-axis represents the outcome scale. The lines represent the trajectories of anxiety for each treatment arm over time. The plot highlights the nature of treatment effects on anxiety as captured by the linear mixed-effects model, allowing for the incorporation of random effects and accounting for within-subject correlations.

### 3.4. Anxiety

The interaction between Study Week and Sleep A (Arm 1) showed a significant negative association with stress (β = -0.360, p = 0.004). This indicates that the effect of Study Week on stress levels differed between the treatment groups, with participants in Sleep A (Arm 1) showing a larger decrease in stress over time compared to the placebo group (Figure 4, Table 5). Additionally, Sex demonstrated a significant negative association with stress (β = -0.666, p = 0.019), suggesting that males reported higher levels of stress compared to females. Age showed a significant negative association with stress (β = -0.065, p < 0.001), indicating that older participants reported lower levels of stress. Furthermore, BMI exhibited a significant positive association with stress (β = 0.033, p = 0.049), indicating that higher BMI was associated with higher stress levels.

**Table 5.** Significant Factors Associated with Stress: Results from a Linear Mixed Effects Regression Model. Summary of significant variables and their associations with stress based on a linear mixed-effects regression model. The model was used to assess the differences in the change in the variables of interest between each active product arm versus placebo. The table presents the beta coefficients (β), standard errors (Std.Error), degrees of freedom (DF), t-values, and p-values for each variable. Higher values indicate a stronger positive association with stress, while lower values indicate a stronger negative association.

|  | Value | Std.Error | DF | t-value | p-value |
| --- | --- | --- | --- | --- | --- |
| (Intercept) | 14.746 | 0.782 | 835 | 18.856 | <0.001 |
| Education: No College Degree [Ref: College Degree] | 0.435 | 0.272 | 610 | 1.597 | 0.111 |
| Sex at Birth: Male [Ref: Female] | -0.666 | 0.283 | 610 | -2.353 | 0.019 |
| Age | -0.065 | 0.012 | 610 | -5.239 | <0.001 |
| Race: Non-white [Ref: white] | -0.287 | 0.352 | 610 | -0.814 | 0.416 |
| Race: Native Hawaiian or Pacific Islander [Ref: white] | -0.108 | 2.331 | 610 | -0.046 | 0.963 |
| BMI | 0.033 | 0.017 | 610 | 1.971 | 0.049 |
| Hispanic, LatinX, or Spanish origin: Yes [Ref: No] | -0.107 | 0.471 | 610 | -0.228 | 0.820 |
| Study Week | -0.521 | 0.088 | 835 | -5.913 | <0.001 |
| Sleep A (Arm 1) | 0.202 | 0.334 | 610 | 0.605 | 0.545 |
| Sleep B (Arm 2) | 0.501 | 0.332 | 610 | 1.507 | 0.132 |
| Study Week: Sleep A (Arm 1) | -0.360 | 0.126 | 835 | -2.864 | 0.004 |
| Study Week: Sleep B (Arm 2) | -0.047 | 0.125 | 835 | -0.372 | 0.710 |

**Figure 4.**
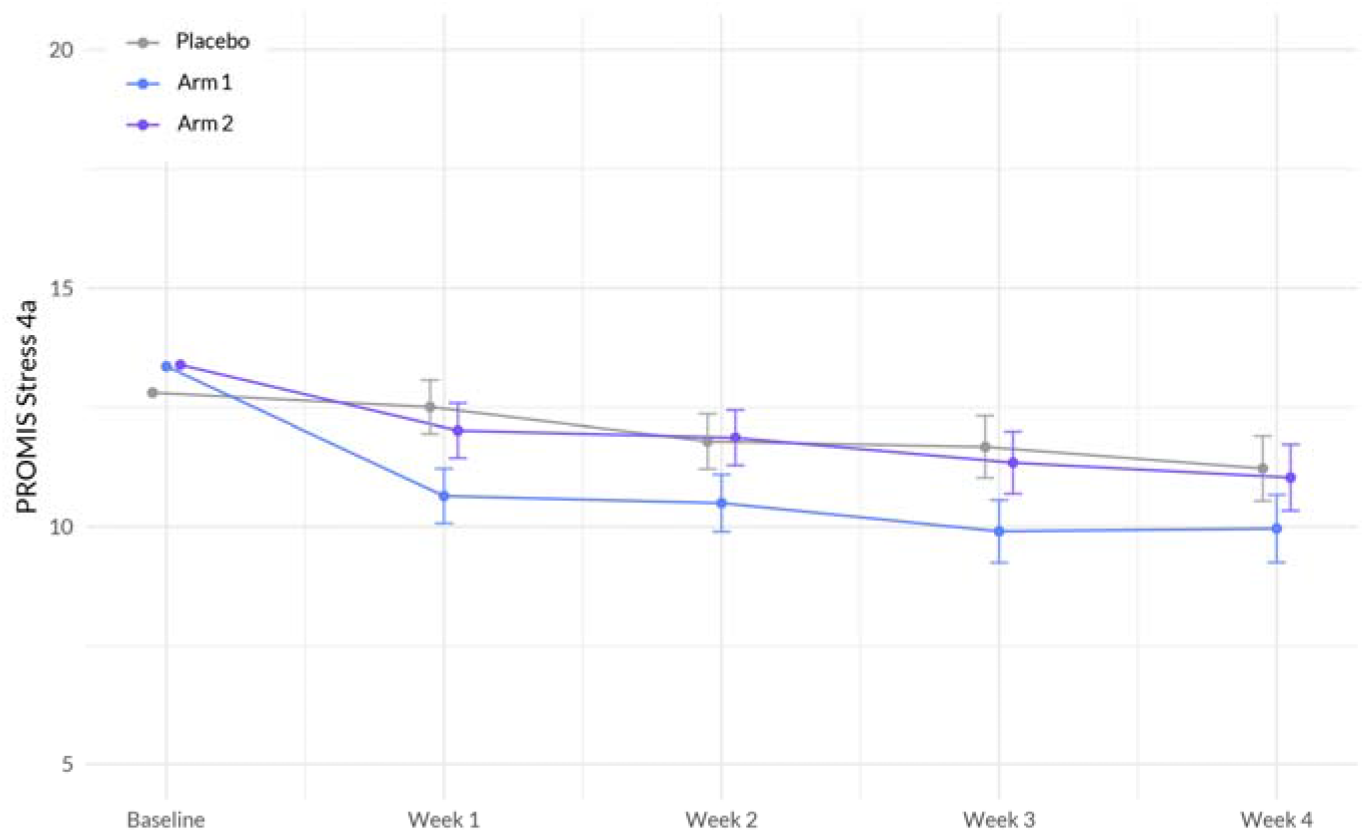
Interaction between Treatment and Week on PROMIS Stress 4a score. The plot illustrates the interaction between treatment (Sleep A; Arm 1 and Sleep B; Arm 2) and week on the stress scale, based on a linear mixed-effects model. The x-axis represents the weeks of the study, while the y-axis represents the outcome scale. The lines represent the trajectories of stress for each treatment arm over time. The plot highlights the nature of treatment effects on stress as captured by the linear mixed-effects model, allowing for the incorporation of random effects and accounting for within-subject correlations.

### 3.5. Pain

Our primary analyses revealed no significant differences in the rate of mean PEG (Pain, Enjoyment, General Activity) score change between Sleep A and placebo (β = -0.024, p = 0.788), or between Sleep B and placebo (β = 0.032, p = 0.713), see Table 6. However, age showed a significant positive association with pain (β = 0.024, p = 0.019), indicating that as age increased, participants reported higher levels of pain. Education also demonstrated a significant positive association with pain (β = 0.706, p = 0.0023), suggesting that individuals with a higher level of education experienced higher levels of pain. Additionally, Study Week showed a significant negative association with pain (β = -0.307, p < 0.001), indicating that as the study progressed, participants reported lower levels of pain.

**Table 6.**
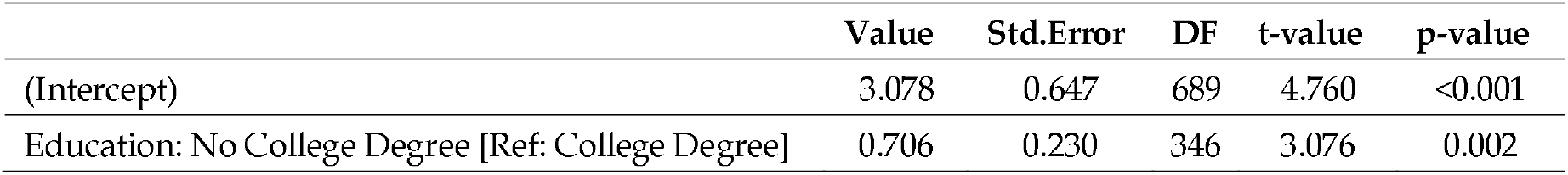
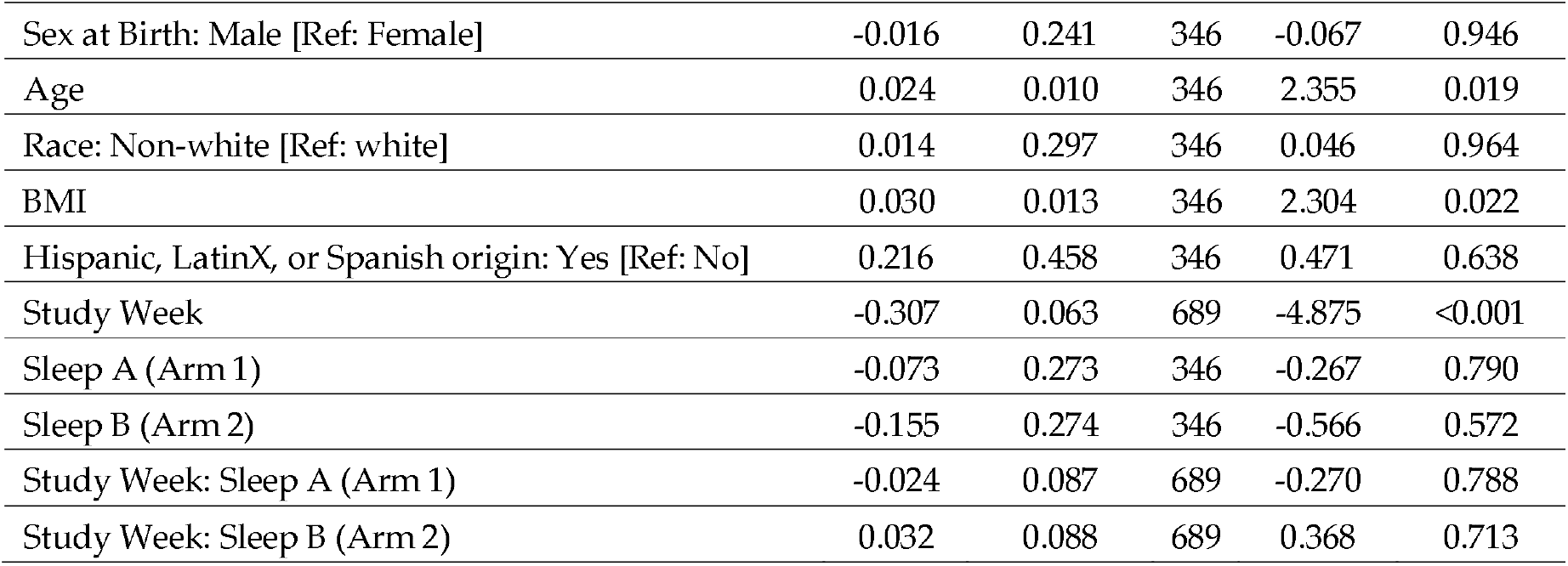
Significant Factors Associated with Pain: Results from a Linear Mixed Effects Regression Model. Summary of significant variables and their associations with pain based on a linear mixed-effects regression model. The model was used to assess the differences in the change in the variables of interest between each active product arm versus placebo. The table presents the beta coefficients (β), standard errors (Std.Error), degrees of freedom (DF), t-values, and p-values for each variable. Higher values indicate a stronger positive association with pain, while lower values indicate a stronger negative association.

### 3.6. Overall well-being

The interaction between Study Week and Sleep A (Arm 1) showed a significant positive association with wellbeing (β = 0.318, p = 0.0346). This indicates that the effect of Study Week on overall wellbeing differed between the treatment groups, with participants in Sleep A (Arm 1) experiencing a greater improvement in wellbeing over time compared to the placebo group. Education demonstrated a significant negative association with wellbeing (β = -1.559, p < 0.001), suggesting that individuals without a college degree reported lower levels of overall wellbeing. BMI exhibited a significant negative association with wellbeing (β = -0.060, p = 0.0061), indicating that higher BMI was associated with lower levels of wellbeing (Figure 5, Table 7).

**Table 7.** Significant Factors Associated with Overall Well-Being: Results from a Linear Mixed Effects Regression Model. Summary of significant variables and their associations with overall well-being based on a linear mixed-effects regression model. The model was used to assess the differences in the change in the variables of interest between each active product arm versus placebo. The table presents the beta coefficients (β), standard errors (Std.Error), degrees of freedom (DF), t-values, and p-values for each variable. Higher values indicate a stronger positive association with overall well-being, while lower values indicate a stronger negative association.

|  | Value | Std.Error | DF | t-value | p-value |
| --- | --- | --- | --- | --- | --- |
| (Intercept) | 11.251 | 1.020 | 1238 | 11.031 | <0.001 |
| Education: No College Degree [Ref: College Degree] | -1.559 | 0.356 | 610 | -4.383 | <0.001 |
| Sex at Birth: Male [Ref: Female] | 0.315 | 0.368 | 610 | 0.856 | 0.392 |
| Age | 0.015 | 0.016 | 610 | 0.898 | 0.369 |
| Race: Non-white [Ref: white] | 0.781 | 0.458 | 610 | 1.706 | 0.089 |
| Race: Native Hawaiian or Pacific Islander [Ref: white] | 1.657 | 2.918 | 610 | 0.568 | 0.570 |
| BMI | -0.060 | 0.022 | 610 | -2.752 | <0.001 |
| Hispanic, LatinX, or Spanish origin: Yes [Ref: No] | 0.266 | 0.616 | 610 | 0.432 | 0.666 |
| Study Week | 0.672 | 0.106 | 1238 | 6.308 | <0.001 |
| Sleep A (Arm 1) | 0.422 | 0.431 | 610 | 0.980 | 0.327 |
| Sleep B (Arm 2) | 0.084 | 0.427 | 610 | 0.196 | 0.844 |
| Study Week: Sleep A (Arm 1) | 0.318 | 0.151 | 1238 | 2.116 | <0.001 |
| Study Week: Sleep B (Arm 2) | -0.005 | 0.148 | 1238 | -0.037 | 0.971 |

**Figure 5.**
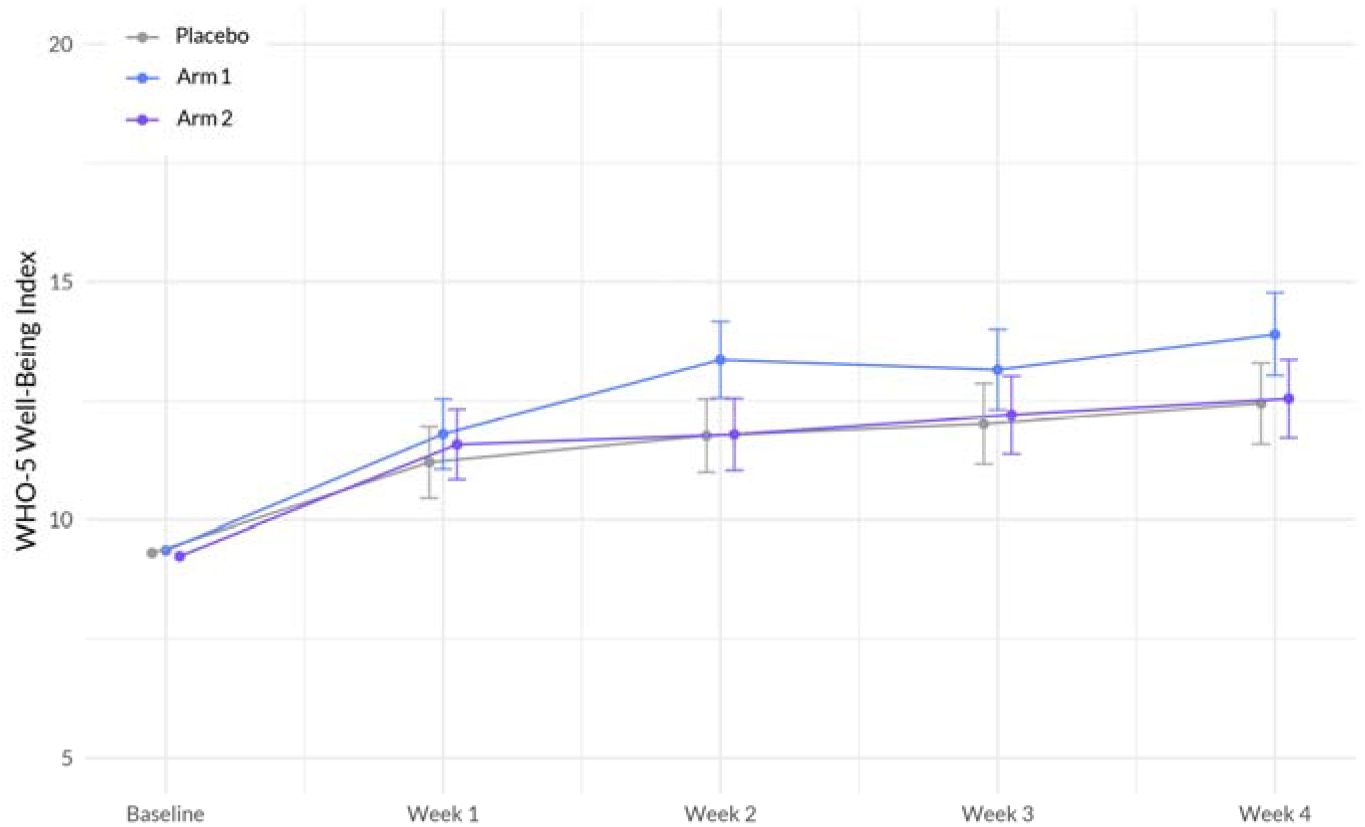
Interaction between Treatment and Week on World Health Organization (WHO)-5 Well-being index. The plot illustrates the interaction between treatment (Sleep A; Arm 1 and Sleep B; Arm 2) and week on the wellbeing scale, based on a linear mixed-effects model. The x-axis represents the weeks of the study, while the y-axis represents the outcome scale. The lines represent the trajectories of wellbeing for each treatment arm over time. The plot highlights the nature of treatment effects on wellbeing as captured by the linear mixed-effects model, allowing for the incorporation of random effects and accounting for within-subject correlations.

### 3.3. Side Effects

Side effects were slightly more common among the active arms (Arm 1 and Arm 2, (χ^2^(2) = 5.64, p = 0.059)), predominantly grogginess and drowsiness and mostly mild; none was considered serious or required use of emergency or non-emergency healthcare services (Figure 6).

**Figure 6.**
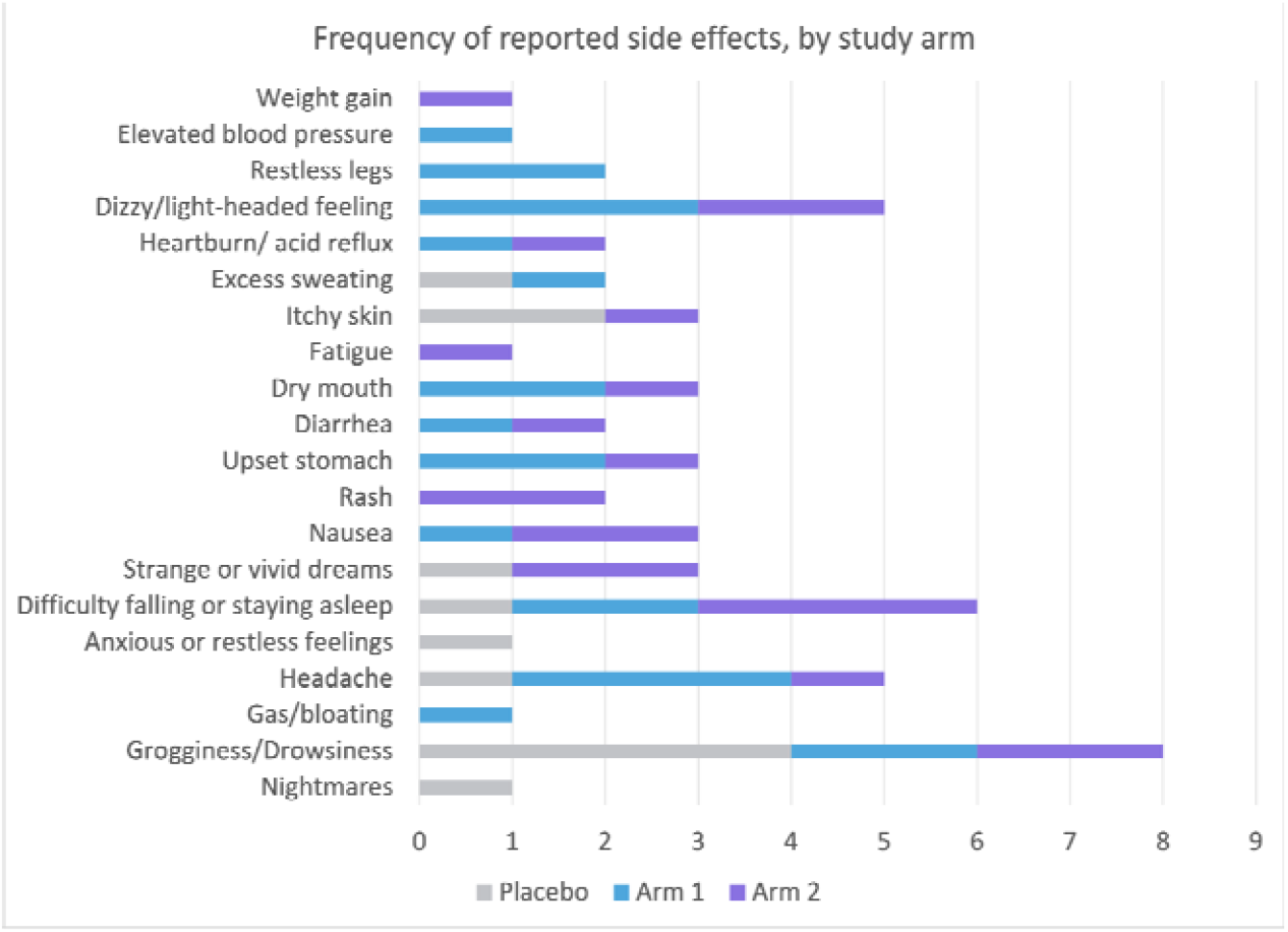
Comparison of side effects between active arms (Sleep A; Arm 1 and Sleep B; Arm 2) and placebo. This figure displays the occurrence of side effects in the active arms (Sleep A; Arm 1 and Sleep B; Arm 2) compared to the placebo group. Participants with sleep disturbance received Sleep A (Arm 1), Sleep B (Arm 2), or placebo for 4 weeks. Side effects, primarily grogginess and drowsiness, were slightly more common in the active arms but were mild and non-serious, requiring no emergency or non-emergency healthcare services.

## 4. Discussion

We presented a randomized, double-blinded, placebo-controlled trial to evaluate the effectiveness and safety of two orally ingested softgel dietary supplements, Sleep A and Sleep B, compared to a placebo over four weeks of treatment. We observed a significant difference in effect on four health outcomes (sleep disturbance, anxiety, stress, and overall well-being) between Sleep A formula and placebo control. We observed no significant difference in effect on any health outcomes between Sleep B formula and placebo control. Importantly, both supplements exhibited favorable safety profiles, as all side effects were mild or moderate and there were no significant differences in the frequency of reported side effects between the active and placebo arms.

This study is among the first to investigate the effectiveness and safety of supplements that aim to enhance sleep and overall health in a large participant sample. These supplements were specifically designed to harness the potential synergistic effects of botanical ingredients that have demonstrated favorable outcomes in promoting better sleep. Our primary analyses indicate that over the course of four weeks using the study product, the Sleep A group experienced a significantly greater reduction in sleep disturbance compared to the placebo group. Additionally, participants in the Sleep A group also showed significantly greater improvements in anxiety, stress, and overall well-being compared to those in the placebo group. These results are expected, given the intricate connections between sleep, anxiety, stress, and overall well-being [34].

Interestingly, we observed no significant differences in any of the health outcomes between Sleep B and placebo. It is important to note that both Sleep A and B formulas contained the same amount of CBD, CBN, and L-Theanine, while Sleep formula A contained a lower amount of THC and higher amounts of GABA, hops oil, and valerian oil than the Sleep B formula. Since the current study did not employ a factorial design to examine the individual effects of all ingredients, as well as their potential interactions, we are unable to determine the main driver(s) between the different impacts Sleep A and Sleep B had on behavioral outcomes. We put forward a hypothesis regarding the potential reasons for the superior performance of Sleep A in reducing sleep disturbances compared to Sleep B. There are two primary factors that we believe contribute to this outcome. Firstly, Sleep A contains a lower amount of THC compared to Sleep B. One study found that those participants who use cannabis multiple times a week for its sleeping effects prefer strains with lower amounts of THC, suggesting that the strains with lower THC may be more effective at promoting sleep [35]. Secondly, Sleep A contains higher levels of three specific ingredients: GABA, hops oil, and valerian oil. These ingredients are known to function as positive modulators of GABA, a neurotransmitter associated with promoting sleep and inducing mild sedation [23,36]. Therefore, it is plausible that the presence of these ingredients in Sleep A contributes to its effectiveness in improving sleep quality.

The data obtained in the present study are consistent with previous findings that a combination of cannabinoids could improve sleep. For example, a randomized, controlled crossover trial administering a combination product, containing THC 20 mg/mL, CBN 2 mg/mL, CBD 1 mg/mL, and naturally occurring terpenes (extracted from the cannabis plant), in pharmaceutical grade sunflower oil, for 2 weeks, demonstrated an improvement in sleep quality in subjects with insomnia when compared to placebo [13]. Similarly, subjects receiving a tablet containing 10 mg THC and 5 mg CBN nightly experienced significantly improved sleep quality and slept significantly longer, with a 5% increase in sleep duration [12]. We recently published the results of a similar sleep study on 1793 adults [37]. Participants were randomly assigned to take 1 of 6 products, containing either 15 mg CBD or 5 mg melatonin, alone or in combination with minor cannabinoids, including CBN. Most participants (56% to 75%) across all formulations experienced a clinically important improvement in their sleep quality though not statistically better than the active control group that took 5 mg of melatonin alone.

This study was intended to approximate the “real world” effectiveness of the study products by administering them to a broad population that used the products in a manner and setting similar to that of actual consumers. Unlike conventional clinical trials, which often have restrictive eligibility criteria and rigorous monitoring, the data in question may exhibit higher levels of missingness and heterogeneity. Nonetheless, conventional clinical trials involving natural products frequently suffer from limited sample sizes and lack external validity, as the participants’ characteristics and behaviors may not accurately represent those of real-world users. Consequently, studies like this try to reflect the real-world effects of such products and possess distinct value in their capacity to provide evidence for regulatory and clinical decision-making and additional clinical trial design [38].

Whether or not the results of our study reflect botanical synergy, meaning that the higher levels of the hops, valerian, and GABA allowed for the lower level of THC to be effective, was not directly investigated in this study. However, it poses interesting questions and warrants further investigation.

This study has multiple limitations. First, approximately 26% of participants did not complete any follow-up surveys. However, the overall attrition level was still below our anticipated attrition (45%) and the study remained adequately powered to detect significant sleep changes. Furthermore, because the products used in this study were combinatorial, we are unable to pinpoint the exact drivers of the observed changes.

Considering the complex combinations of products examined in this study, further investigations are warranted to identify the specific active ingredient(s) responsible for the observed significant effects. To achieve this, a rigorous study employing a factorial design with multiple arms would be beneficial. This design would systematically vary the ingredients in different combinations, allowing for the examination of both their individual effects and potential interactions on behavioral outcomes. Such an approach would enhance our understanding of the precise drivers behind the observed effects.

## 5. Conclusions

In this randomized, double-blind, placebo-controlled trial to evaluate the effects of two formulations of sleep softgels relative to placebo, we observed that a botanical blend containing lower amounts of THC and higher amounts of GABA, hops oil, and valerian oil significantly improved sleep quality, anxiety, stress, and overall well-being in healthy individuals with a desire for better sleep. We observed no significant difference in effect on any health outcome (sleep quality, anxiety, stress, pain, or overall well-being) between a botanical blend containing higher amounts of THC, and lower amounts of GABA, hops oil, and valerian oil and the placebo control. The active products demonstrated a favorable safety profile; all side effects were mild or moderate, and there were no significant differences in the frequency of reported side effects between the active arms and placebo.

## Data Availability

All data produced in the present study are available upon reasonable request to the authors

## Author Contributions

Conceptualization and Methodology, J. Chen and E. Pauli. Data curation and formal analysis, C. Bryant.; writing—original draft preparation, A. Kolobaric and S.Hewlings.; writing—review and editing, all authors. All authors have read and agreed to the published version of the manuscript.”

## Institutional Review Board Statement

The study was conducted in accordance with the Declaration of Helsinki, and approved by Sterling Independent Review Board (SIRB) [10147]. The master protocol Radicle Rest™ was registered on ClinicalTrials.gov [Identifier: NCT05511818]. It should be noted that the protocol registered is not this specific study but rather is the templated study protocol utilized for all Radicle Rest studies.

## Informed Consent Statement

Informed consent was obtained from all subjects involved in the study.

## Data Availability Statement

The data presented in this study are available on request from the corresponding author. The data are not publicly available.

## Acknowledgments

The authors would like to express our deepest gratitude to the participant volunteers in this study without whom this study would not have been possible. We acknowledge and appreciate the hard work of the Radicle Science study operations team and MDbio The Doctors Brand™ for the product formulas studied.

## Disclaimer/Publisher’s Note

The statements, opinions and data contained in all publications are solely those of the individual author(s) and contributor(s) and not of MDPI and/or the editor(s). MDPI and/or the editor(s) disclaim responsibility for any injury to people or property resulting from any ideas, methods, instructions or products referred to in the content.

## References

1. Altman, N.G.; Izci-Balserak, B.; Schopfer, E.; Jackson, N.; Rattanaumpawan, P.; Gehrman, P.R.; Patel, N.P.; Grandner, M.A. Sleep duration versus sleep insufficiency as predictors of cardiometabolic health outcomes. Sleep medicine 2012, 13, 1261–1270, doi:10.1016/j.sleep.2012.08.005.

2. Golem, D.L.; Martin-Biggers, J.T.; Koenings, M.M.; Davis, K.F.; Byrd-Bredbenner, C. An integrative review of sleep for nutrition professionals. Advances in nutrition (Bethesda, Md.) 2014, 5, 742–759, doi:10.3945/an.114.006809.

3. McCoy, J.G.; Strecker, R.E. The cognitive cost of sleep lost. Neurobiology of learning and memory 2011, 96, 564–582, doi:10.1016/j.nlm.2011.07.004.

4. Institute of Medicine Committee on Sleep, M.; Research. The National Academies Collection: Reports funded by National Institutes of Health. In Sleep Disorders and Sleep Deprivation: An Unmet Public Health Problem, Colten, H.R., Altevogt, B.M., Eds.; National Academies Press (US) Copyright © 2006, National Academy of Sciences.: Washington (DC), 2006.

5. Reynolds, A.C.; Paterson, J.L.; Ferguson, S.A.; Stanley, D.; Wright, K.P., Jr.; Dawson, D. The shift work and health research agenda: Considering changes in gut microbiota as a pathway linking shift work, sleep loss and circadian misalignment, and metabolic disease. Sleep medicine reviews 2017, 34, 3–9, doi:10.1016/j.smrv.2016.06.009.

6. Doherty, R.; Madigan, S.; Warrington, G.; Ellis, J. Sleep and Nutrition Interactions: Implications for Athletes. Nutrients 2019, 11, doi:10.3390/nu11040822.

7. Verma, K.; Singh, D.; Srivastava, A. The Impact of Complementary and Alternative Medicine on Insomnia: A Systematic Review. Cureus 2022, 14, e28425, doi:10.7759/cureus.28425.

8. Babson, K.A.; Sottile, J.; Morabito, D. Cannabis, Cannabinoids, and Sleep: a Review of the Literature. Current psychiatry reports 2017, 19, 23, doi:10.1007/s11920-017-0775-9.

9. Tham, M.; Yilmaz, O.; Alaverdashvili, M.; Kelly, M.E.M.; Denovan-Wright, E.M.; Laprairie, R.B. Allosteric and orthosteric pharmacology of cannabidiol and cannabidiol-dimethylheptyl at the type 1 and type 2 cannabinoid receptors. British journal of pharmacology 2019, 176, 1455–1469, doi:10.1111/bph.14440.

10. Suraev, A.S.; Marshall, N.S.; Vandrey, R.; McCartney, D.; Benson, M.J.; McGregor, I.S.; Grunstein, R.R.; Hoyos, C.M. Cannabinoid therapies in the management of sleep disorders: A systematic review of preclinical and clinical studies. Sleep medicine reviews 2020, 53, 101339, doi:10.1016/j.smrv.2020.101339.

11. Taylor, L.; Gidal, B.; Blakey, G.; Tayo, B.; Morrison, G. A Phase I, Randomized, Double-Blind, Placebo-Controlled, Single Ascending Dose, Multiple Dose, and Food Effect Trial of the Safety, Tolerability and Pharmacokinetics of Highly Purified Cannabidiol in Healthy Subjects. CNS drugs 2018, 32, 1053–1067, doi:10.1007/s40263-018-0578-5.

12. Gannon, W.; Bronfein, W.; Jackson, D.; Holshouser, K.; Artman, B.E.; Schestepol, M.; Treacy, D.J.; Rudnic, E.M. Novel Formulation of THC and CBN in a Repeat-Action Tablet Improves Objective and Subjective Measurements of Sleep. Am J Endocannabinoid Medicine 2021, 3, 12–18.

13. Walsh, J.H.; Maddison, K.J.; Rankin, T.; Murray, K.; McArdle, N.; Ree, M.J.; Hillman, D.R.; Eastwood, P.R. Treating insomnia symptoms with medicinal cannabis: a randomized, crossover trial of the efficacy of a cannabinoid medicine compared with placebo. Sleep 2021, 44, doi:10.1093/sleep/zsab149.

14. Corroon, J. Cannabinol and Sleep: Separating Fact from Fiction. Cannabis and cannabinoid research 2021, 6, 366–371, doi:10.1089/can.2021.0006.

15. 15. Gottesmann, C. GABA mechanisms and sleep. Neuroscience 2002, 111, 231–239, doi:10.1016/s0306-4522(02)00034-9.

16. Ngo, D.H.; Vo, T.S. An Updated Review on Pharmaceutical Properties of Gamma-Aminobutyric Acid. Molecules (Basel, Switzerland) 2019, 24, doi:10.3390/molecules24152678.

17. Saeed, M.; Naveed, M.; Arif, M.; Kakar, M.U.; Manzoor, R.; Abd El-Hack, M.E.; Alagawany, M.; Tiwari, R.; Khandia, R.; Munjal, A.; et al. Green tea (Camellia sinensis) and l-theanine: Medicinal values and beneficial applications in humans-A comprehensive review. Biomedicine & pharmacotherapy = Biomedecine & pharmacotherapie 2017, 95, 1260–1275, doi:10.1016/j.biopha.2017.09.024.

18. Kim, S.; Jo, K.; Hong, K.B.; Han, S.H.; Suh, H.J. GABA and l-theanine mixture decreases sleep latency and improves NREM sleep. Pharmaceutical biology 2019, 57, 65–73, doi:10.1080/13880209.2018.1557698.

19. Ives, A. An experimental inquiry in the chemical properties and economical and medicinal virtues of the Humulus lupulus, or common hop. Ann. Philos 1821, 2, 194–202.

20. Biendl, M.; Pinzl, C. Hops and health. MBAA TQ 2009, 46, 1–7.

21. Nuutinen, T. Medicinal properties of terpenes found in Cannabis sativa and Humulus lupulus. European journal of medicinal chemistry 2018, 157, 198–228.

22. Franco, L.; Sánchez, C.; Bravo, R.; Rodriguez, A.; Barriga, C.; Juánez, J.C. The sedative effects of hops (Humulus lupulus), a component of beer, on the activity/rest rhythm. Acta physiologica Hungarica 2012, 99, 133–139, doi:10.1556/APhysiol.99.2012.2.6.

23. Benkherouf, A.Y.; Soini, S.L.; Stompor, M.; Uusi-Oukari, M. Positive allosteric modulation of native and recombinant GABA(A) receptors by hops prenylflavonoids. European journal of pharmacology 2019, 852, 34–41, doi:10.1016/j.ejphar.2019.02.034.

24. Carbone, K.; Gervasi, F. An Updated Review of the Genus Humulus: A Valuable Source of Bioactive Compounds for Health and Disease Prevention. Plants (Basel, Switzerland) 2022, 11, doi:10.3390/plants11243434.

25. Xu, H.; Yuan, H.; Pan, L.; Guo, X. The Pharmacological effects of volatile oil from Valeriana on central nervous system. Chinese Journal of Pharmaceutical Analysis 1997, 17, 399–400.

26. Li, J.; Li, X.; Wang, C.; Zhang, M.; Ye, M.; Wang, Q. The potential of Valeriana as a traditional Chinese medicine: Traditional clinical applications, bioactivities, and phytochemistry. Frontiers in Pharmacology 2022, 13.

27. Orhan, I.E. A Review Focused on Molecular Mechanisms of Anxiolytic Effect of Valerina officinalis L. in Connection with Its Phytochemistry through in vitro/in vivo Studies. Current pharmaceutical design 2021, 27, 3084–3090, doi:10.2174/1381612827666210119105254.

28. Bonn-Miller, M.O.; ElSohly, M.A.; Loflin, M.J.E.; Chandra, S.; Vandrey, R. Cannabis and cannabinoid drug development: evaluating botanical versus single molecule approaches. International review of psychiatry (Abingdon, England) 2018, 30, 277–284, doi:10.1080/09540261.2018.1474730.

29. Efferth, T.; Koch, E. Complex interactions between phytochemicals.The multi-target therapeutic concept of phytotherapy. Current drug targets 2011, 12, 122–132, doi:10.2174/138945011793591626.

30. Wagner, H.; Ulrich-Merzenich, G. Synergy research: approaching a new generation of phytopharmaceuticals. Phytomedicine : international journal of phytotherapy and phytopharmacology 2009, 16, 97–110, doi:10.1016/j.phymed.2008.12.018.

31. Russo, E.B. Taming THC: potential cannabis synergy and phytocannabinoid-terpenoid entourage effects. British journal of pharmacology 2011, 163, 1344–1364, doi:10.1111/j.1476-5381.2011.01238.x.

32. Norman, G.R.; Sloan, J.A.; Wyrwich, K.W. Interpretation of changes in health-related quality of life: the remarkable universality of half a standard deviation. Medical care 2003, 41, 582–592, doi:10.1097/01.Mlr.0000062554.74615.4c.

33. Yu, L.; Buysse, D.J.; Germain, A.; Moul, D.E.; Stover, A.; Dodds, N.E.; Johnston, K.L.; Pilkonis, P.A. Development of short forms from the PROMIS™ sleep disturbance and sleep-related impairment item banks. Behavioral sleep medicine 2012, 10, 6–24.

34. Alvaro, P.K.; Roberts, R.M.; Harris, J.K. A Systematic Review Assessing Bidirectionality between Sleep Disturbances, Anxiety, and Depression. Sleep 2013, 36, 1059–1068, doi:10.5665/sleep.2810.

35. Belendiuk, K.A.; Babson, K.A.; Vandrey, R.; Bonn-Miller, M.O. Cannabis species and cannabinoid concentration preference among sleep-disturbed medicinal cannabis users. Addictive behaviors 2015, 50, 178–181, doi:10.1016/j.addbeh.2015.06.032.

36. Granger, R.E.; Campbell, E.L.; Johnston, G.A. (+)- And (-)-borneol: efficacious positive modulators of GABA action at human recombinant alpha1beta2gamma2L GABA(A) receptors. Biochemical pharmacology 2005, 69, 1101–1111, doi:10.1016/j.bcp.2005.01.002.

37. Saleska, J.L.; Bryant, C.; Kolobaric, A.; D’Adamo, C.R.; Colwell, C.S.; Loewy, D.; Chen, J.; Pauli, E.K. The Safety and Comparative Effectiveness of Non-Psychoactive Cannabinoid Formulations for the Improvement of Sleep: A Double-Blinded, Randomized Controlled Trial. Journal of the American Nutrition Association 2023, 1–11, doi:10.1080/27697061.2023.2203221.

38. Sherman, R.E.; Anderson, S.A.; Dal Pan, G.J.; Gray, G.W.; Gross, T.; Hunter, N.L.; LaVange, L.; Marinac-Dabic, D.; Marks, P.W.; Robb, M.A.; et al. Real-World Evidence - What Is It and What Can It Tell Us? The New England journal of medicine 2016, 375, 2293–2297, doi:10.1056/NEJMsb1609216.

